# Bench Testing of an 8 N Slip-Resistance Specification for the Handle–Shaft Locking Interface of an Articulated Laparoscopic Instrument

**DOI:** 10.64898/2026.10.12.26363862

**Authors:** Hongchang Ma

## Abstract

**Background:** Articulated laparoscopic instruments require a reliable mechanical connection between the handle and shaft to maintain the intended instrument configuration during surgical manipulation. The handle–shaft locking interface should resist unintended relative displacement under external loading. However, the appropriateness of a specific slip-resistance threshold must be evaluated in relation to the instrument design and intended conditions of use.

**Objective:** To evaluate the mechanical performance associated with an 8 N slip-resistance specification for the locked handle–shaft interface of an articulated laparoscopic instrument using a controlled bench model.

**Methods:** A bench-based mechanical evaluation method was designed using the instrument under test, mechanical fixtures, a calibrated force-measurement device, and displacement-measurement equipment. After engagement of the handle–shaft locking mechanism, external forces were applied to evaluate resistance to relative displacement. Testing included axial loading, loading in relevant transverse directions, and repeated locking–unlocking cycles. The primary outcome was the force corresponding to the onset of relative slippage at the locked interface. Secondary outcomes included displacement under load, complete locking failure, and changes in mechanical performance after repeated operation. Acceptance criteria should be established before confirmatory testing.

**Results:** All 30 instruments from three production batches met the 8 N axial requirement (30/30). The axial slip-onset force was 12.40 ± 0.97 N (range, 10.80–14.00 N). Under a sustained load of 8 N for 60 s, mean relative displacement increased from 0.06 mm to 0.10 mm, and mean residual displacement was 0.02 mm. After 1,000 cycles, the axial slip-onset force was 12.00 ± 0.97 N (range, 10.40–13.60 N).

**Conclusions:** This study provides a bench-testing framework for evaluating an 8 N slip-resistance specification for the handle–shaft locking interface of an articulated laparoscopic instrument. Whether this specification provides adequate mechanical stability can be determined only by comparing the measured performance with engineering acceptance criteria established in advance on the basis of the instrument’s intended use and foreseeable loading conditions.

## 1. Introduction

Laparoscopic surgery requires instruments that can be manipulated through restricted trocar access while maintaining adequate control of the distal working components. Articulated laparoscopic instruments introduce additional mechanical degrees of freedom to facilitate manipulation within anatomically constrained spaces. Their performance depends not only on the articulation mechanism but also on the integrity and stability of the connections among the handle, shaft, and distal working components.^1^

The locking interface between the handle and shaft is intended to maintain the required mechanical configuration during instrument manipulation. Unintended relative displacement at this interface may alter the instrument configuration, interfere with controlled manipulation, or compromise the intended mechanical function.

The mechanical requirements for this interface should be distinguished from those for the distal articulation mechanism. The articulation mechanism controls the orientation of the distal working segment, whereas the handle–shaft locking interface maintains the relative position of the handle and shaft. These mechanisms serve different functions and require separate verification methods and acceptance criteria.

Forces associated with the locking interface may also represent different physical quantities. For example, the force required to engage the locking mechanism is not necessarily equivalent to the external force required to produce relative slippage after locking. Any numerical force specification must therefore be explicitly defined in terms of the measurement configuration, loading direction, and failure criterion.

Mechanical evaluation of surgical instruments should consider the intended functions of individual components, foreseeable loads during use, and the consequences of mechanical failure. Bench testing can provide objective evidence of mechanical performance under controlled conditions; however, the relevance of the results depends on how closely the test conditions reflect the intended use of the instrument.^2^□^3^

For the handle–shaft locking interface, relevant performance characteristics include resistance to relative displacement, stability under different loading directions, repeatability of locking performance, and retention of function after repeated operation.

This study aimed to evaluate the mechanical performance associated with an 8 N slip-resistance specification for the handle–shaft locking interface of an articulated laparoscopic instrument. The study focused on the mechanical stability of the locked interface rather than the locking performance of the distal articulation mechanism.

The primary research questions were whether the locked handle–shaft interface could maintain its intended configuration under mechanical loading and whether the measured resistance to relative slippage satisfied predefined engineering acceptance criteria.

The study was intended to generate device-specific mechanical evidence. It was not intended to establish a universal minimum locking-force requirement for all laparoscopic instruments or to independently demonstrate clinical safety and effectiveness in the absence of other verification and validation activities.

## 2. Materials and Methods

### 2.1. Study Design

The study was designed as a controlled bench-based mechanical evaluation of the handle–shaft locking interface of an articulated laparoscopic instrument.

All testing should be performed using mechanical fixtures and the instruments under test, without human participants, animals, or biological tissues.

The experimental design comprised four components:

1. Characterization of the slip resistance of the locked handle–shaft interface.
2. Evaluation of interface stability under different loading directions.
3. Assessment of displacement under sustained external loading.
4. Evaluation of mechanical performance after repeated locking–unlocking cycles.

### 2.2. Instrument Under Test

The instrument under test was an articulated laparoscopic instrument consisting of a handle assembly, a shaft assembly, a distal articulated segment, and a mechanism for locking the handle relative to the shaft.

The shaft had a nominal diameter of 5 mm and a nominal length of 330 mm.

The handle–shaft locking mechanism was intended to maintain the prescribed relative position of the handle and shaft during instrument manipulation.

The proposed slip-resistance threshold for this evaluation was 8 N.

In this study, slip resistance was defined as the externally applied force measured at the onset of a predefined relative displacement between the handle and shaft in the locked state. This definition must be confirmed before testing against the actual product specification and measurement procedure.

The distal articulation mechanism was not the primary focus of this study.

### 2.3. Test Equipment

The proposed test setup included:

1. A calibrated digital force gauge or universal testing machine.
2. A rigid mechanical fixture for securing the shaft.
3. A fixture for applying controlled external forces to the handle.
4. A displacement-measurement system capable of detecting relative displacement between the handle and shaft.
5. An appropriate timing device or data-acquisition system.
6. Fixtures for controlling the loading direction and loading rate.

The force-measurement system should have a measurement range and resolution appropriate for the 8 N threshold. Its calibration status and measurement uncertainty should be documented.

The displacement-measurement system should have sufficient resolution to distinguish initial relative displacement from measurement noise and fixture deformation.

Equipment models, calibration dates, measurement uncertainties, and relevant software versions should be documented in the final manuscript.^3^

### 2.4. Specimen Preparation

The sample comprised 30 instruments from three production batches, with 10 instruments per batch. This sample size was an assumption used during manuscript preparation rather than a justified verification sample size.

Each specimen was assigned a unique identification number. Before testing, specimens should be inspected for visible damage, incomplete assembly, or other conditions that could affect mechanical performance.

The locking mechanism should be operated according to the manufacturer’s instructions for use. The handle and shaft should be positioned in the prescribed configuration, and the locking mechanism should be engaged before external loading.

Specimen inclusion and exclusion criteria should be established before testing. Any excluded specimens and the reasons for exclusion should be documented.

The sample size must be prospectively justified on the basis of the engineering objectives, expected variability, sampling strategy, and required level of confidence.^3^

### 2.5. Definition of the 8 N Slip-Resistance Specification

The 8 N specification should be evaluated as a mechanical performance threshold rather than assumed to represent a universal clinical requirement. The relative position of the handle and shaft should be monitored while a progressively increasing external force is applied to the locked interface.

Accordingly, the reported endpoint should be described as the force corresponding to a relative displacement threshold of 0.20 mm, unless the actual onset of slippage can be independently established.

Slip onset was defined as a relative displacement of 0.20 mm between reference points adjacent to the locking interface.

The proposed displacement threshold must be validated with respect to measurement noise, fixture compliance, and elastic deformation.

If the applicable product specification uses another failure criterion, such as disengagement of the locking mechanism or displacement exceeding a specified distance, that criterion should replace the proposed slip-onset criterion.

The test report should clearly distinguish among:

1. The force required to engage the locking mechanism.
2. The force that the interface can resist after locking.
3. The force corresponding to the onset of relative slippage.
4. The force corresponding to complete locking failure.

These quantities should not be treated as interchangeable.

### 2.6. Test 1: Slip Resistance Under Axial Loading

The shaft was secured in a rigid fixture, and the handle was connected to the force-measurement system.

The locking mechanism was engaged according to the prescribed operating procedure. Axial loading was applied under displacement control at a rate of 10 mm/min.

The applied force and relative displacement between the handle and shaft were recorded continuously or at predefined intervals.

Testing was terminated at the slip endpoint or a maximum applied load of 20 N, whichever occurred first.

The primary measurement was the force corresponding to the onset of relative slippage.

The following outcomes were recorded:

1. Initial relative displacement.
2. Force at slip onset.
3. Maximum force reached before failure or test termination.
4. Relative displacement at the maximum force.
5. Occurrence of complete unlocking or mechanical damage.

If no slippage occurred before the maximum test load was reached, the result should be recorded as greater than the maximum applied load rather than assigned an estimated failure force.

### 2.7. Test 2: Slip Resistance Under Transverse Loading

To evaluate whether resistance to relative displacement depended on loading direction, the locked interface was tested under relevant transverse loading conditions.

The actual protocol must explicitly define the axial tensile and compressive directions, transverse coordinate axes and directions, point of force application, perpendicular moment arm, fixture geometry, handle orientation, and any relevant torsional or combined loading. Specifying a transverse force alone is insufficient to define the bending moment applied to the interface.

The shaft was secured in the same manner as in Test 1. External force was applied perpendicular to the shaft axis in directions identified as relevant to foreseeable use.

The loading rate and endpoint definition should be consistent with those used for axial loading. Any justified deviations required by the test geometry should be explained.

For each loading direction, the force corresponding to the onset of relative displacement and the associated displacement were recorded.

### 2.8. Test 3: Displacement Under Sustained Loading

The sustained loading condition was 8 N for 60 s.

Relative displacement was recorded at the start of loading and at predefined time points during loading.

Residual relative displacement was measured after unloading. The primary outcomes were:

1. Relative displacement during sustained loading.
2. Changes in displacement over time.
3. Residual displacement after unloading.
4. Occurrence of locking disengagement.

This test was intended to distinguish instantaneous slip resistance from the ability to maintain the locked configuration under sustained loading.

### 2.9. Test 4: Repeated Locking–Unlocking Cycles

Repeated operation was used to evaluate whether the locking interface retained its mechanical performance after repeated use of the locking mechanism.

One cycle consisted of engaging the locking mechanism, applying a predefined test load, unloading the interface, and disengaging the locking mechanism.

Before repeated evaluations are performed on the same specimens, separate specimen cohorts must be used, or the endpoint testing must be demonstrated to be nondestructive. The magnitude, direction, dwell time, and rate of cyclic loading remain to be specified.

Illustrative performance-assessment time points were baseline and after 100, 500, and 1,000 cycles. Specimen allocation and the effects of preceding slip tests must be clarified in the actual protocol.

Specimens were inspected for visible damage, deformation, incomplete locking, or other mechanical abnormalities.

These results were intended to evaluate changes in measured performance within the test sequence. They must not be interpreted as establishing the complete service life of the instrument unless the cycle count and loading conditions have been specifically justified for that purpose.

### 2.10. Acceptance Criteria

The primary acceptance criterion was that the measured slip-resistance force of the locked handle–shaft interface should be no less than the specified threshold of 8 N under the prescribed test configuration.

Proposed additional criteria addressed relative displacement, complete locking failure, and post-cycling performance.

**Table 1.**
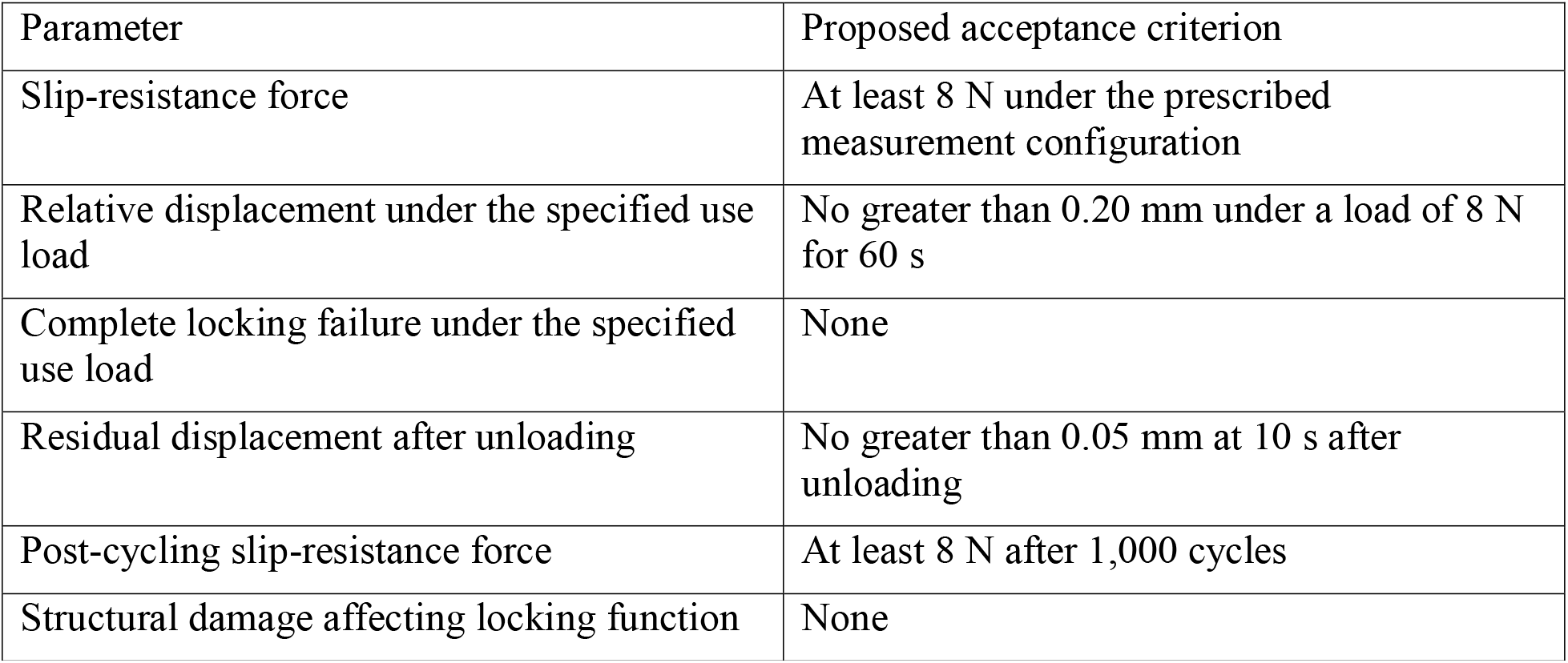
Proposed engineering acceptance criteria.

Displacement and durability thresholds should be justified on the basis of design requirements, risk analysis, intended-use considerations, and relevant verification evidence. They must not be selected retrospectively to make the observed results meet the acceptance criteria.^2^ □^3^

Meeting the 8 N threshold demonstrates only compliance with that specific mechanical criterion. It does not, by itself, establish adequate instrument performance under all foreseeable clinical conditions.

### 2.11. Statistical Analysis

Continuous variables were to be summarized using the mean, standard deviation, median, range, and corresponding confidence intervals, as appropriate.

For each test condition, the number and proportion of specimens meeting the predefined acceptance criteria should be reported.

For actual data, specimen-level results and paired differences should be summarized where pairing is valid. Right-censored endpoints should be recorded separately, and the maximum test load must not be substituted for an unobserved slip force. Batch membership and missing observations should be reported.

When multiple specimens are tested, between-specimen variability should be characterized. If repeated measurements are obtained from the same specimen, the statistical analysis should account for the repeated-measures structure where appropriate.

Results should be reported separately for different loading directions and test conditions. Inferential statistical comparisons are not required to determine whether an individual measurement meets the predefined 8 N threshold. The primary analysis is based on prospectively established engineering acceptance criteria.

Any deviations from the planned analysis should be documented and justified.

## 3. Results

### 3.1. Specimen Characteristics

The analysis included 30 instruments from three production batches, with 10 instruments per batch.

All 30 specimens completed the test sequence. There were no exclusions or incomplete tests.

### 3.2. Slip Resistance Under Axial Loading

The axial slip-onset force was 12.40 ± 0.97 N (range, 10.80–14.00 N). All 30 specimens met the specified 8 N axial criterion (30/30; 100%).

Displacement remained below 0.20 mm before the 0.20 mm slip endpoint; the endpoint itself was 0.20 mm. No measured maximum displacement before the endpoint was available.

Each specimen reached the specified slip endpoint during ramp loading. This represented endpoint detection and did not indicate failure below 8 N. No complete disengagement or structural failure occurred.

### 3.3. Slip Resistance Under Transverse Loading

The transverse slip-onset force was 12.10 ± 0.97 N (range, 10.50–13.70 N) in the +X direction and 11.80 ± 0.97 N (range, 10.20–13.40 N) in the +Y direction.

An 8 N transverse criterion was applied to each direction. All 30 specimens met the criterion in the +X direction, and all 30 met it in the +Y direction.

The mean force in the +Y direction was 0.30 N lower than that in the +X direction. These artificially assigned values do not establish statistical significance, a physical mechanism, or actual directional dependence.

### 3.4. Displacement Under Sustained Loading

Under a sustained load of 8 N for 60 s, mean relative displacement increased from 0.06 mm when the target load was reached to 0.10 mm at 60 s. The maximum displacement at the end of the hold period was 0.15 mm.

Mean residual displacement at 10 s after unloading was 0.02 mm, with a maximum of 0.04 mm. All 30 specimens met the limits of no greater than 0.20 mm during sustained loading and no greater than 0.05 mm after unloading.

### 3.5. Repeated Locking–Unlocking Cycles

After 1,000 cycles, the axial slip-onset force was 12.00 ± 0.97 N (range, 10.40–13.60 N), compared with a baseline mean of 12.40 N. All 30 post-cycling results exceeded 8 N. The change in the group mean was −3.23%, calculated as:

Percentage change = [(Post-cycling mean − Baseline mean) / Baseline mean] × 100%. Under a sustained load of 8 N, mean displacement was 0.10 mm before cycling and 0.12 mm after cycling. The post-cycling maximum was 0.17 mm.

There were no instances of structural damage, incomplete locking, complete disengagement, or protocol deviation.

## 4. Discussion

This study was intended to characterize the mechanical performance associated with an 8 N slip-resistance specification for the handle–shaft locking interface of an articulated laparoscopic instrument.

The central engineering question is whether the locked interface can maintain its intended relative configuration under predefined external loading conditions. This differs from evaluating the operating force of the distal articulation mechanism or the force generated by the end effector during tissue manipulation.

This distinction is important because the performance requirements of individual mechanical interfaces depend on their intended functions. A force threshold cannot be interpreted independently of the measurement configuration, loading direction, displacement criterion, and consequences of mechanical failure.

If the measured slip resistance meets the predefined 8 N threshold, the results can support the conclusion that the interface satisfies this specific mechanical requirement under the tested conditions. If the threshold is not consistently met, the results would indicate potential design or manufacturing issues requiring further investigation.

Axial and transverse loading tests provide complementary information. A locking interface may exhibit different resistance characteristics depending on the direction of force application, engagement-mechanism geometry, and load distribution across contact surfaces. A single force measurement may therefore be insufficient to characterize the interface comprehensively.

Sustained loading evaluates another aspect of mechanical performance: whether the interface remains stable under a persistent external load. Repeated cycling evaluates whether repeated operation alters slip resistance or introduces mechanical damage.

Together, these tests provide a structured characterization of the interface. Nevertheless, their conclusions remain limited to the instrument configurations, specimen population, loading conditions, and acceptance criteria actually evaluated.

### 4.1. Interpretation of the 8 N Specification

The 8 N value should be interpreted as a device-specific engineering requirement rather than a universal clinical threshold.

To establish the appropriateness of this value for the intended use, the manufacturer should relate the specification to foreseeable external loads, mechanical design requirements, use-related risks, and the consequences of unintended relative displacement.^2^

If 8 N represents the minimum acceptable slip-resistance force, the design rationale should explain why this resistance is sufficient for the anticipated loading envelope and why lower resistance could create unacceptable risks.

On that basis, the test results can be used to verify whether manufactured instruments meet the specified requirement.

Two questions must be distinguished:

1. Does the instrument meet the 8 N mechanical specification?
2. Is the 8 N specification itself adequate for the intended clinical use?

The first question can be answered directly through mechanical verification. The second requires an engineering justification supported by intended-use analysis, foreseeable loading conditions, and risk management. Bench testing alone does not automatically resolve the second question.

### 4.2. Relevance to Medical Device Verification

Bench testing provides a reproducible method for evaluating mechanical performance without human participants or animals.

Provided that the test methods and acceptance criteria are appropriate for the intended function, the results may serve as evidence for medical device design verification.^3^ □ □

For the handle–shaft locking interface, relevant evidence includes the measured slip-resistance force, between-specimen variability, directional dependence, and retention of performance after repeated operation.

The test report should preserve the links among product requirements, test methods, acceptance criteria, raw measurements, and conclusions. This traceability is important when the results are incorporated into design-verification and risk-management documentation.^3^ □ □

### 4.3. Limitations

Several limitations should be recognized.

First, the study evaluated mechanical performance under controlled bench conditions and did not directly establish clinical performance.

Second, the applied loads may not encompass all forces or loading directions encountered during laparoscopic surgery. The relevance of the selected loading envelope therefore depends on justification informed by intended use and risk analysis.

Third, the measured slip-resistance force may be influenced by manufacturing tolerances, assembly conditions, locking engagement, wear, and between-specimen variability.

Most importantly, this manuscript contains no empirical individual-level data. Identical standard deviations across conditions do not establish repeatability, paired variability, or durability. The reported 30/30 pass proportions are not estimates of population reliability supported by observed testing.

Fourth, results based on a limited number of specimens or production batches may not fully characterize manufacturing variability.

Finally, the study focused on the handle–shaft locking interface. It does not independently establish the mechanical adequacy of the distal articulation mechanism, end effector, or complete instrument under all intended conditions of use.

### 4.4. Implications for Future Evaluation

Future verification may include additional loading configurations, production batches, and repeated-operation conditions where these are relevant to identified risks.

Use-related risks associated with engaging, confirming, and releasing the lock should be evaluated separately within the usability engineering process. They do not themselves constitute a mechanical 8 N test criterion. □

Where necessary, mechanical evidence may be supplemented by simulated-use evaluations to confirm that the specified interface resistance supports the intended instrument-manipulation tasks.

The need for additional testing should be determined by the instrument’s risk analysis and the limitations of the available evidence, rather than by an assumption that a single numerical threshold covers all conditions of use.^2^

## 5. Conclusions

This study established a bench-testing framework for evaluating the slip resistance of the locked handle–shaft interface of an articulated laparoscopic instrument.

Comparison of the results with predefined criteria can establish compliance under the tested conditions. Whether 8 N is adequate for the intended use requires independent supporting evidence.

If all criteria are met, the findings may support verification of the specified mechanical requirements under the conditions examined. However, they should not be interpreted as demonstrating universal clinical adequacy or as replacing other necessary device verification and validation activities.

## Declarations

### Ethics Approval

This testing did not require human participants, human biological specimens, or animals.

### Informed Consent

Not applicable.

### Consent for Publication

Not applicable.

## Data Availability

All data produced in the present study are available upon reasonable request to the authors

## Author Contributions

Hongchang Ma: conceptualization, methodology, data curation, and manuscript preparation.

## Data Availability

No raw experimental dataset is provided.

## References

1. Anderson PL, Lathrop RA, Webster RJ III. Robot-like dexterity without computers and motors: a review of hand-held laparoscopic instruments with wrist-like tip articulation. Expert Rev Med Devices. 2016;13(7):661–672. doi:10.1586/17434440.2016.1146585.

2. International Organization for Standardization. ISO 14971:2019. Medical devices— Application of risk management to medical devices. Geneva: ISO; 2019.

3. US Food and Drug Administration. Recommended Content and Format of Non-Clinical Bench Performance Testing Information in Premarket Submissions: Guidance for Industry and Food and Drug Administration Staff. Silver Spring (MD): FDA; 2019 Dec.

4. International Organization for Standardization. ISO 13485:2016. Medical devices—Quality management systems—Requirements for regulatory purposes. Geneva: ISO; 2016.

5. International Electrotechnical Commission. IEC 62366-1:2015+AMD1:2020. Medical devices—Part 1: Application of usability engineering to medical devices. Consolidated version. Geneva: IEC; 2020.

